# The Cambridge Centre for Ageing and Neuroscience (Cam-CAN) longitudinal study protocol: Phase 4 (“Enrichment”) and Phase 5 (“Rescan”)

**DOI:** 10.1101/2025.05.06.25327023

**Authors:** Ina Demetriou, Adam Attaheri, Tina Bingham, William Duckett, Lara Bridge, Petar Raykov, Kamen A. Tsvetanov, Marta Correia, Dace Apšvalka, Maite Crespo-Garcia, Karen Campbell, Alexa Morcom, Daniel J. Mitchell, James Rowe, Noham Wolpe, Sarah E. Henderson, Cam-CAN, Richard Henson

## Abstract

**Background:** The Cambridge Centre for Ageing and Neuroscience (Cam-CAN) started in 2010 to study the effect of healthy adult ageing on cognition and the brain in a population-derived sample. The study design and protocol for Phases 1-3 of Cam-CAN were detailed in Shafto et al. (2014); this paper outlines the design and protocol of Phases 4–5, which enable longitudinal investigation of cognitive and brain ageing over approximately 12 years. More details about the Cam-CAN project can be found here: www.cam-can.org.

**Methods/Design:** Phase 4 was an at-home assessment of cognition, demographics and lifestyle, performed approximately 6 years after Phase 1 (baseline assessment), for which all people from Phase 1 were invited. Phase 5 combined repeated online cognitive, demographics and lifestyle assessment, followed by in-lab attendance for MRI and MEG brain scanning, approximately 12 years after Phase 1, for which all people from Phase 2 (baseline brain assessment) were invited. Demographics, lifestyle and cognitive data are therefore now available for three timepoints, and MRI and MEG brain data for two timepoints.

**Discussion:** The Cam-CAN study offers deep and wide phenotyping of neurocognitive health across the adult lifespan (18-96). These rich data will allow researchers to address questions like: why do some people maintain their cognitive abilities better than others, in terms of their brain structure or function, their lifestyle and/or their genetics? Given the shifting demographics towards old age in most countries, this knowledge will be important to help people function independently for longer, reducing both individual and societal burden.

## Background

People typically live longer than they did in previous generations, and the proportion of the population that is in “old age” (typically seen as over 65 years old) has been steadily increasing in most countries. Ageing brings cognitive decline, owing to brain changes, and these problems have costs both on individuals and the society that cares for them. Therefore, a better understanding of how the brain ages, and how this affects cognition, is important for designing social policy and interventions, e.g. in lifestyles, that will allow people to maintain their cognitive abilities for longer. This is particularly pressing owing to the rapidly-changing environments that people inhabit, which are becoming more cognitively challenging, e.g., where many daily activities depend increasingly on internet services, mobile phones, etc.

The Cambridge Centre for Ageing and Neuroscience (CamCAN) (https://cam-can.mrc-cbu.cam.ac.uk) was established in 2010 to address such questions, by studying the cognitive neuroscience of healthy ageing. CamCAN complements other cohorts related to ageing, brain and cognition, such as the UK Biobank (https://www.ukbiobank.ac.uk), CFAS and CC75 studies (https://www.phpc.cam.ac.uk), the 1946 Birth Cohort and Lothian Birth Cohorts (https://lothian-birth-cohorts.ed.ac.uk). In comparison to these other initiatives, CamCAN’s protocol is distinguished by (i) the population-base, with recruitment aimed at representing the community from which participants are drawn; (ii) the even-distribution of ages, across the adult lifespan; and (iii) the intensity of cognitive and imaging assessments, including extensive and multi-modal functional brain imaging.

CamCAN is a virtual centre, including several departments within the University of Cambridge (see www.cam-can.org). It was founded by an award from the UK Biotechnology and Biological Sciences Research Council (BBSRC), which enabled recruitment of a sample of healthy individuals across the adult lifespan who contributed data on their lifestyle, cognitive abilities and brain imaging, the latter obtained using magnetic resonance imaging (MRI) and magnetoencephalography (MEG). Full details of the initial CamCAN protocol, for Phases (Stages) 1-3^1^, can be found in Shafto et al. (2014). To date, the CamCAN data have been used in over 240 peer-reviewed scientific publications, enabled via a minimal data use agreement (available from: https://camcan-archive.mrc-cbu.cam.ac.uk/dataaccess).

There is increasing appreciation of the importance of longitudinal data to study ageing. While it has long been known that cross-sectional data on brain and cognition are confounded by generational differences in birth year (e.g., changes in lifestyle, nutrition, education over the years), recent comparison of cross-sectional versus longitudinal effects of age have revealed that important factors like education affect baseline cognitive abilities and brain volumes in old age, but not changes in those abilities/volumes over time (Nyberg et al., 2021, Fjell et al., 2025).

Furthermore, predictions of cross-sectional age from brain images do not reliably predict longitudinal brain change (Vidal-Pineiro et al., 2021; see Walhovd et al., 2024, for review). This motivated the longitudinal assessment of the CamCAN cohort, with support from the European Union Horizon 2020 scheme (specifically the LifeBrain project, https://www.lifebrain.uio.no) and intramural funding from the Medical Research Council Cognition & Brain sciences Unit (MRC CBU). More specifically, we conducted two further phases to obtain longitudinal lifestyle and cognitive data (Phase 4) and most recently, repeat MRI and MEG (Phase 5). The repeat brain data, over an interval of approximately 12 years on average, is particularly valuable to study neurocognitive ageing. A timeline for the CamCAN project is shown in Figure 1. This paper describes the nature of the recruitment and data collection from Phases 4-5.

**Figure 1.**
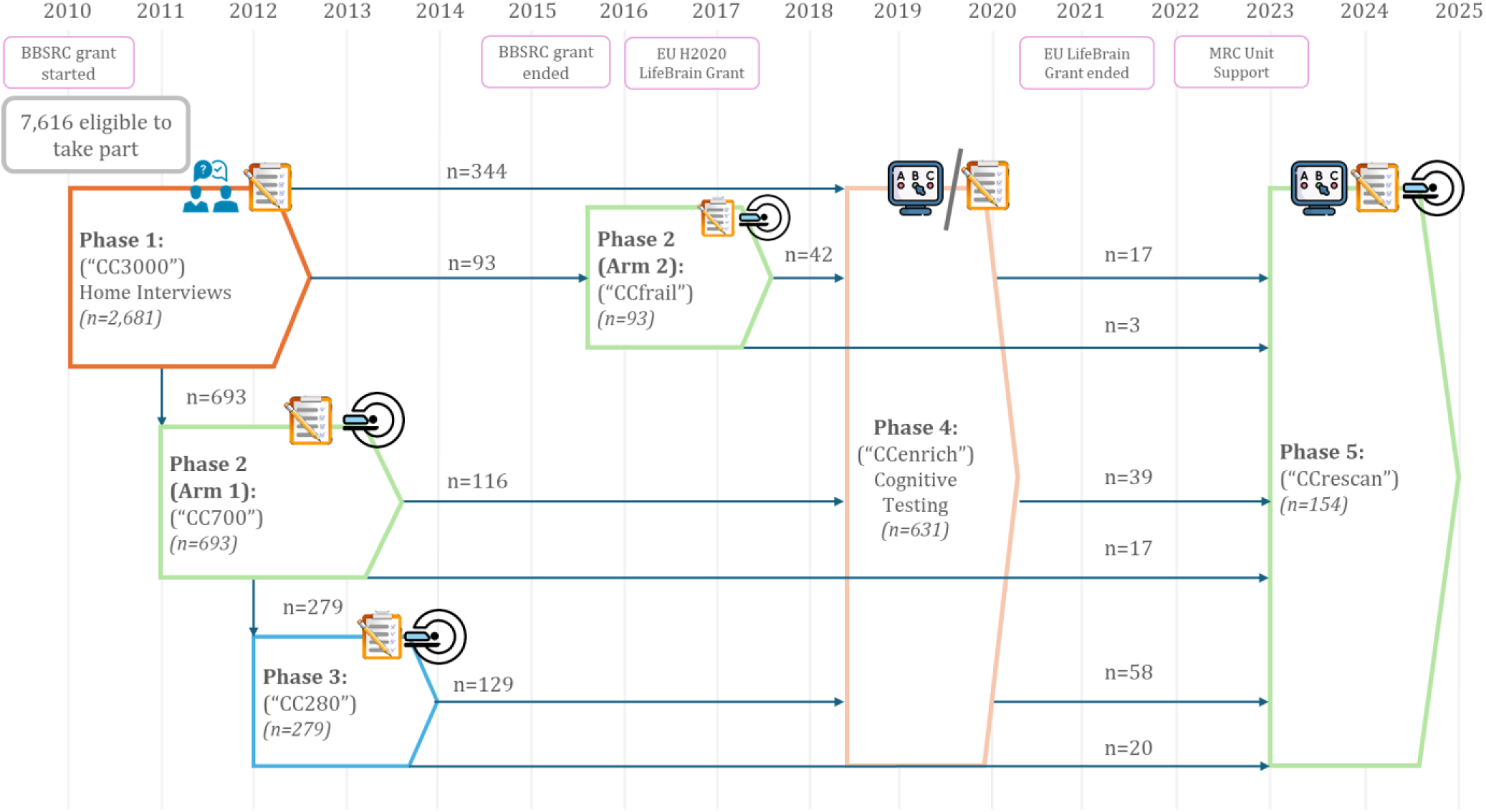
CamCAN study participation timeline with corresponding number of participants retained from each phase. Green/Blue – neuroimaging data available. Orange – cognitive data only, of varying formats. Pen and paper icon – paper testing (in-lab/home), computer icon – home online testing, MRI icon – neuroimaging data available (MRI & MEG), blue people icon – home interview. Years of phases are approximated (+/- 1 year deviations from the graph) – for precise years of data collection – refer to the main text.

## Summary of previous Cam-CAN Phases 1-3

Potential participants were identified from General Practitioner (GP) surgeries in Cambridge, United Kingdom. In the UK, 98% of citizens are registered with a GP, for provision of primary care services. In the CamCAN catchment area, a total of 7,616 individuals (see Green et al., 2018) were eligible and approached, with follow-up home visits for non-responders. Of these, 2,681 (35.2 %) agreed to participate in the study.

In Phase 1 of the study (2011-13; also called the “CC3000” sample), participants completed a 2- hour home interview to obtain basic demographic, lifestyle, health, and cognitive data.

In Phase 2-Arm 1 (overlapping 2011-13; also called the “CC700” sample), approximately 100 participants per decade (ages 18–88 years; spanning seven decades) were randomly selected from Phase 1, provided they met health criteria for brain imaging and had no history of dementia, referral for dementia assessment, nor memory complaints, and had a Mini-Mental State Examination (MMSE) score greater than 24 (Folstein et al., 1975) (see Shafto et al., 2014, for details). A total of 708 participants (Arm1 and Arm2 together) completed at least one visit to the MRC CBU. These visits included specialised cognitive tests as well as 3T Magnetic Resonance Imaging (MRI) and 306-channel Magnetoencephalography (MEG) sessions for brain imaging.

In Phase 3 (2013-14; also called “CC280” sample), a subset of 280 participants from Phase 2- Arm1 were recalled to the CBU for three additional visits, including further MRI and MEG sessions with more specialised cognitive tasks. Full details of the tests and brain imaging epidemiological characteristics of the sample can be found in Green et al. (2018). Some of the scientific outcomes of Phases 1-3 are available here: https://cam-can.mrc-cbu.cam.ac.uk/publications/.

In Phase 2-Arm 2 (2016-18; also called “CCfrail” sample), a second independent sample from Phase 1 underwent a similar cognitive and brain imaging protocol to Phase 2-Arm1. This sample included 42 participants who scored below the MMSE cut-off of 24 for dementia but had not reported cognitive problems to their GP, plus a further 51 age- and sex-matched controls who passed the MMSE cut-off (as well as patients with Mild Cognitive Impairment recruited independently from local Memory Clinics, who are not part of CamCAN, see Kocagoncu et al., 2022).

## Cam-CAN Phase 4: Enrichment

### Design

Phase 4 was conducted from 2018-19 as an ‘Enrichment Phase’ (“CCenrich”), with two main aims: 1) to collect repeat data on selected lifestyle, health, and cognitive variables, and 2) to collect new data on variables not previously recorded, but harmonised with those from other European cohorts in the LifeBrain consortium (https://www.lifebrain.uio.no). Due to funding constraints, testing was limited to self-administered, at-home assessments, completed either via posted paper forms, or online through a dedicated website. A total of 1,778 people from Phase 1 were invited (contact had been lost with the remaining 903), and 631 completed at least one cognitive test, including 287 who had also participated in Phase 2. All participants completed the ‘core’ tasks, which were available in both paper and online formats. A subset of participants also completed a selection of ‘extended’ tasks, if they wished, which were available only online (Table 1). The tasks are detailed below. Participants were reimbursed with a £10 Amazon voucher for completing the core tasks, with an additional £10 for completing extended tasks. Participants could not refuse the payment, but when it was not wanted, it was suggested that they donate to a charity themselves.

**Table 1.** Phase 4 (Enrichment) session content and approximate timing, listed by category.

| **Measure** | **Modality** | **Format** | **~ Time (mins)** |
| --- | --- | --- | --- |
| ***Core*** |  |  |  |
| The Science of Ageing Questionnaire | Questionnaire | Online/Paper | 20 |
| The Cattell Test | Cognitive | Online/Paper | 20 |
| Spot the Word Task | Cognitive | Online/Paper | 5 |
| ***Extended*** |  |  |  |
| Reaction Time Task | Cognitive | Online | 8 |
| Digit Span (number recall task) | Cognitive | Online | 10 |
| Spatial Span (location recall task) | Cognitive | Online | 10 |
| Digit-Letter Substitution Task | Cognitive | Online | 2 |
| Story Memory Task | Cognitive | Online | 10 |
| Emotional Stroop (Interference Task) | Cognitive | Online | 10 |
| Stop-Signal, Go/No-Go | Cognitive | Online | 30 |
| Images of Ageing Scale (IAS) | Questionnaire | Online | 10 |
| The Lifetime Depression Assessment Self-Report (LIDAS) | Questionnaire | Online | 15 |
| Eysenck Personality Questionnaire (EPQ) | Questionnaire | Online | 20 |

### Core Tasks

#### The Science of Ageing Questionnaire

A self-completed questionnaire covering core demographics, education, early life experiences, health and well-being, memory (10MQ), current psychological health (assessed using the Hospital Anxiety and Depression Scale, HADS), history of psychological health, family history, social contacts, masculinity and femininity, women’s health, and lifestyle and physical activity. A PDF of the postal versions from Phase 1, Phase 4 and Phase 5 can be found here https://cam-can.mrc-cbu.cam.ac.uk/materials/, along with a CSV file indicating which specific questions appeared in each of Phases 1, 4 and 5.

#### The Cattell Test (fluid intelligence)

The ability to solve novel tasks, often referred to as fluid intelligence, is a core metric of healthy ageing and declines with age. To assess fluid intelligence, participants completed the “Cattell” test (Cattell & Cattell, 1960), specifically Scale 2 Form A of the Cattell Culture Fair test, which was also used in Phase 2 (Shafto et al., 2014). The test consisted of four subtests (series completion, classification, matrices, and topological judgements), each containing progressively more difficult nonverbal pattern-matching puzzles. Each puzzle presented five multiple-choice options. The four blocks were strictly timed to 3 minutes, 4 minutes, 3 minutes, and 2.5 minutes respectively. Participants could choose to complete the test either online, or via a postal pack. Traditionally, the Cattell test is administered by the researcher and the participants are unaware of the timings. However, for the self-administered version in this phase, if performed on paper, participants were instructed how to time themselves correctly, and if performed online, the timing was controlled by the computer. In both formats, participants were aware of the exact timings. There were 46 puzzles in total across the blocks, with the total score being the sum of correct puzzles. The reliability of this test, and any effects of test format (paper vs online), are reported in Demetriou et al. (2024). The same task was used in Phase 2 (in person, standard paper and pencil administration) and Phase 5 (online).

#### Spot the Word task (crystallised intelligence)

Crystallised intelligence, in contrast to fluid intelligence, represents general knowledge about the world and does not decline in healthy ageing (Horn & Cattell, 1967). To assess crystallised intelligence, we used the Spot the Word task from The Speed and Capacity of Language- Processing (SCOLP) assessment (Baddeley et al., 1993). Participants were presented with 60 pairs of items, each containing one real word and one plausible pseudoword, and were asked to identify the real word. Before the main task, they completed a 6-pair practice session with feedback on their responses. The frequency of real words occurring in the English language decreases across trials, making later trials harder. The time to complete the task was unlimited. Participants were encouraged to guess the answer if they were unsure. There was no option to skip an item pair when done online. The total score was the sum of correct trials. The same task was used in Phase 1 (administered in-lab with paper and pencil) and Phase 5 (administered online).

### Extended Tasks

#### Reaction Time Task (processing speed)

‘Simple’ and ‘choice’ reaction time (RT) tasks were used to assess processing speed. Simple RT was based on the time to respond to a single visual stimulus, while choice RT was based on the time to respond to one of four stimuli. Participants were presented with a picture of the right hand on the screen. In the simple RT block, a black dot appeared above only the index finger at varying intervals, following a positively skewed pseudo-random distribution (minimum: 1.8s, mean: 3.7s, median: 3.9s, maximum: 6.8s). Participants were asked to press a key on the keyboard with their right index finger as soon as they saw the black dot. The dot disappeared upon response, triggering the next inter-trial interval (ITI) or disappearing automatically after 3 seconds. This block included 50 trials. In the choice RT block, participants were asked to place four fingers of their right hand on four adjacent keys and respond as quickly as possible to a black dot that appeared above any finger, by pressing the corresponding key. Timing parameters were the same as in the simple RT version, and there were 67 trials in total. The same task was used in Phase 1 (administered at-home on a laptop) and Phase 5 (administered online).

#### Digit Span (verbal working memory)

To assess verbal short-term and working memory, participants completed forward and backward Digit Span tasks, respectively. In both tasks, a sequence of numbers appeared on the screen, one at a time. After the sequence finished, participants responded by selecting the numbers on a keypad. In the forward span task, they had to enter numbers in the same order that they saw them; in the backward span task, they had to enter the numbers in the reverse order. Each task started with three practice trials with feedback provided. In the forward task, participants completed three trials per sequence length from 3 to 13 digits, with the task stopping after three consecutive incorrect responses. For the backward task, the sequence length started at two digits instead. The score for both tasks was the longest sequence that could be recalled correctly at least once.

#### Spatial Span (spatial working memory)

The spatial span task followed the same structure as the digit span task, but instead of numbers, participants were presented with grey boxes on the screen. One box at a time briefly flashed blue in a specific sequence. Participants were asked to remember the sequence of spatial locations. In the forward version, participants clicked the grey boxes in the same order they had flashed. In the backward version, they clicked the boxes in reverse order. All task parameters and scoring were identical to the digit span task.

#### Digit-Letter Substitution Task (processing speed)

This was an online adaptation of the traditional pen-and-paper Digit-Letter Substitution task from the Wechsler Adult Intelligence Scale – Revised (WAIS-R) Manual (Wechsler, 1987).

Participants were presented with a key that matches letters to digits. The task was to use the key to fill in each of the empty boxes with the digit that corresponds to the letter above it. A correctly filled table was given as an example. Participants first practiced with 6 digit-letter pairs and could not proceed until the practice was completed correctly. The main task consisted of 125 digit- letter pairs and was limited to 2 minutes. Participants were unaware the task was timed but were encouraged to complete it as quickly and accurately as possible (finishing all 125 items within 2 minutes was virtually impossible). Participants were instructed to not correct any mistakes and had to continue without revising previous responses. The final score was the total number of correctly substituted digits.

#### Story Memory Task (verbal episodic memory)

To avoid interference from previous administration of the “story memory” task from Phase 1 (which was Story A of the “logical memory” test of the Wechsler Memory Scale III edition; Wechsler (1997), a new story was created that was approximately matched in style, number of units of information and word count. Participants read the story via sequential presentation of small meaningful units, which appeared on the screen at fixed intervals. They were instructed to read each unit carefully and remember it exactly as written. Immediately after the story finished, they were asked to type out everything they could remember (“immediate recall”). After approximately 15 minutes, during which they performed the Stroop task (see below), they were asked to type out the remembered story again (“delayed recall”). Performance was scored by the number of meaningful units recalled, scoring criteria described in Wechsler (1997). The same task was used in Phase 1 and Phase 5 (both administered in person using paper and pencil), but Phase 4 and Phase 5 did not include the third test of delayed recognition.

#### Emotional Stroop (inhibitory control)

Participants completed a standard Stroop task (Stroop, 1935) followed by an Emotional Stroop task (Williams et al., 1996) to assess their ability to inhibit cognitive interference from incongruent stimuli. In the standard Stroop task, participants were presented with one of three words: “RED”, “BLUE”, or “GREEN”, displayed in red, blue, or green ink. The task was to identify the ink colour by pressing the ‘r’ key for red, ‘b’ for blue, or ‘g’ for green. The task began with 24 practice trials, evenly split between 12 congruent trials (where the word and ink colour matched) and 12 incongruent trials (where they did not match). Feedback was provided during the practice phase. This was followed by the main task, which consisted of 96 trials (48 congruent and 48 incongruent) with no feedback. If a participant did not respond within 1500 milliseconds, the task automatically progressed to the next trial.

In the Emotional Stroop task, participants were presented with pictures of either happy or sad unidentifiable. A word was superimposed in the centre of each image, drawn from either a ‘sad’ category (e.g. gloomy, upset, miserable), or a ‘happy’ category (e.g. cheerful, joyful, glad).

Participants were asked to identify the emotional category of the words while ignoring expressions of the faces. The task began with 15 practice trials (4 congruent, 6 incongruent, 5 neutral) with feedback provided. The main task consisted of 96 trials (32 congruent, 32 incongruent, 32 neutral), with no feedback. The main outcome measure was the difference between reaction times and errors on congruent vs. incongruent trials.

#### Stop-Signal, Go/No-Go (inhibitory control)

This task assessed response inhibition, the ability to suppress an initiated or prepotent action. It combined elements of the standard Stop-Signal and Go/No-Go paradigms. The task began with a series of practice trials, before the main experiment. The experiment started with a one-minute practice, followed by the main experiment, which lasted approximately 18 minutes and was conducted over two blocks. Each trial started with a fixation cross (1,500ms), followed by a Go/No-Go/Stop-Signal stimulus. Across the two blocks, there were 360 “Go” trials, in which participants saw a black arrow either pointing left (180) or right (180) and were asked to press the corresponding left or right key with their right hand. On 80 “Stop-Signal” trials (20 left and 20 right), a black arrow was initially presented but turned red after a variable delay, accompanied by a tone. Participants were instructed to stop their response in these cases. The stop-signal delay (SSD) was initially set at 200ms less than the participant’s average reaction time (RT), estimated from 20 Go trials presented at the start of the experiment. The SSD then dynamically adjusted in 50ms steps to maintain ∼50% successful inhibition, with a minimum of 50ms and a maximum of 950ms. Each trial allowed a maximum response time of 1,000ms. If no response was made within this window, the next trial began immediately. On the 40 “No-Go” trials (20 left and 20 right), a red arrow appeared, and participants were asked to withhold any response (max duration: 1,000ms). No-Go trials are equivalent to a Stop-trial with SSD=0ms. Trial types were randomised throughout the experiment. The task was divided into two blocks with a short break in between. Key outcome measures included commission error rate on Stop and No-Go trials, mean RT on correct Go trials, and RT for commission error rate in Stop and No-Go trials, and the estimated time to inhibit a response in Stop-Signal trials. A similar version of this task was performed during MEG and MRI sessions in Phase 3, and during the MEG session in Phase 5.

#### Optional Questionnaires

Participants could also optionally complete the following standardised questionnaires: Images of Ageing Scale (IAS) (Levy et al., 2004), The Lifetime Depression Assessment Self-Report (LIDAS) (Bot et al., 2017), and Eysenck Personality Questionnaire (EPQ) (Eysenck & Eysenck, 2011).

## Cam-CAN Phase 5: Rescan

### Design

Phase 5 was conducted from 2023-25 as a ‘Rescan Phase’ (“CCrescan”), with the main aim of obtaining follow-up brain and cognitive data approximately 12 years after the initial brain measures collected in Phase 2. All healthy participants from Phase 2 (both Arm 1 and Arm 2) who had not informed us that they were unable to continue participation were invited to participate in Phase 5. A total of 574 people were contacted (contact had been lost with the remaining 212), and 243 expressed interest to participate after the initial contact. Out of these, 10 were not eligible due to hearing/vision/mobility problems. To date, 154 have completed at least one Phase 5 session, 63 did not respond to repeat invites, the remaining 16 either did not want to participate, moved away from Cambridge, could not be contacted, were too unwell to participate, or had died.

The study included core online tasks plus two in-lab neuroimaging sessions (MRI and MEG) at the CBU. The order of the MRI and MEG sessions was counterbalanced across age tertiles.

Participants were also invited to complete optional extended cognitive tasks. The measures collected during the online and in-lab sessions are listed in Table 2 and described in more detail below.

**Table 2.**
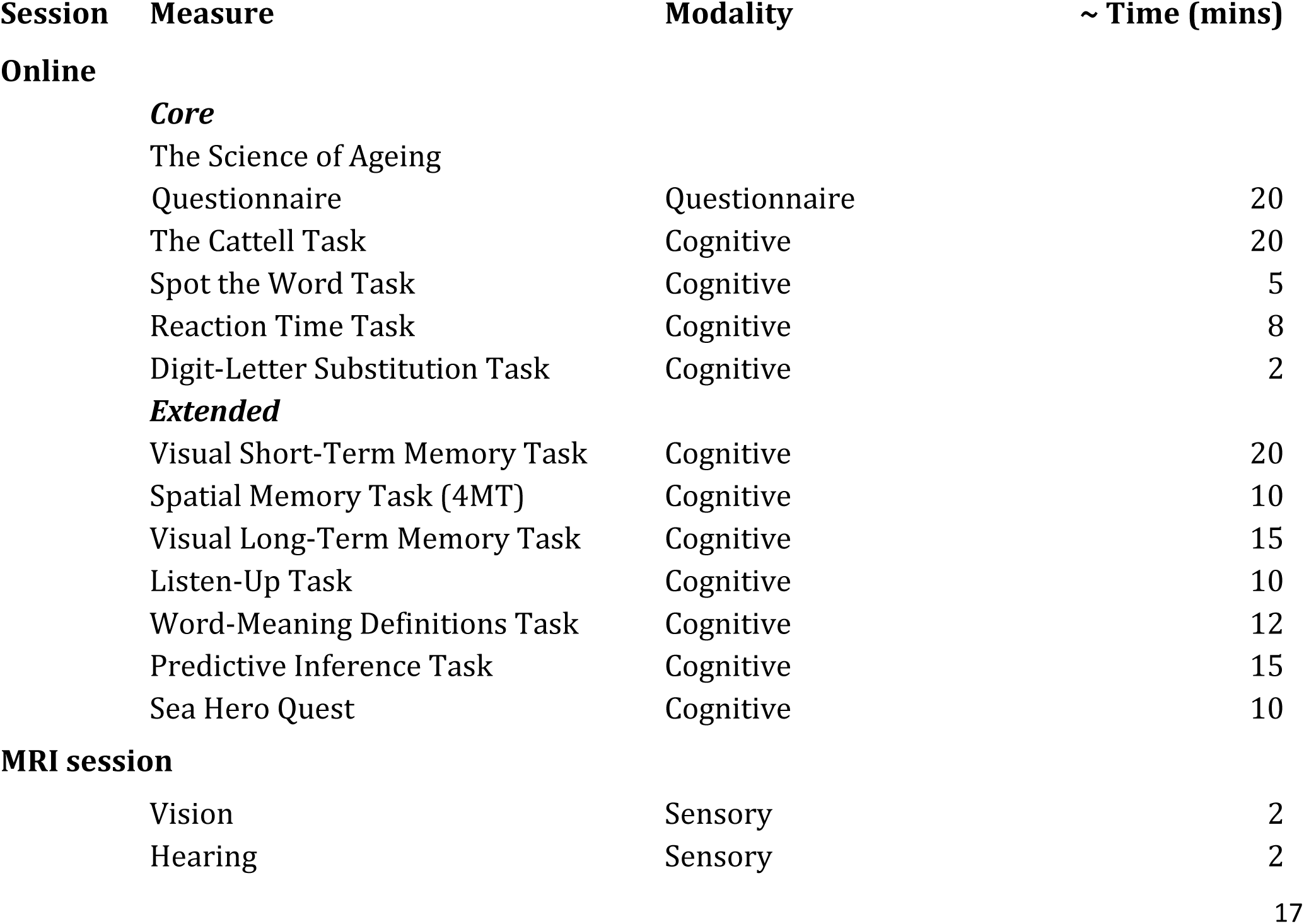
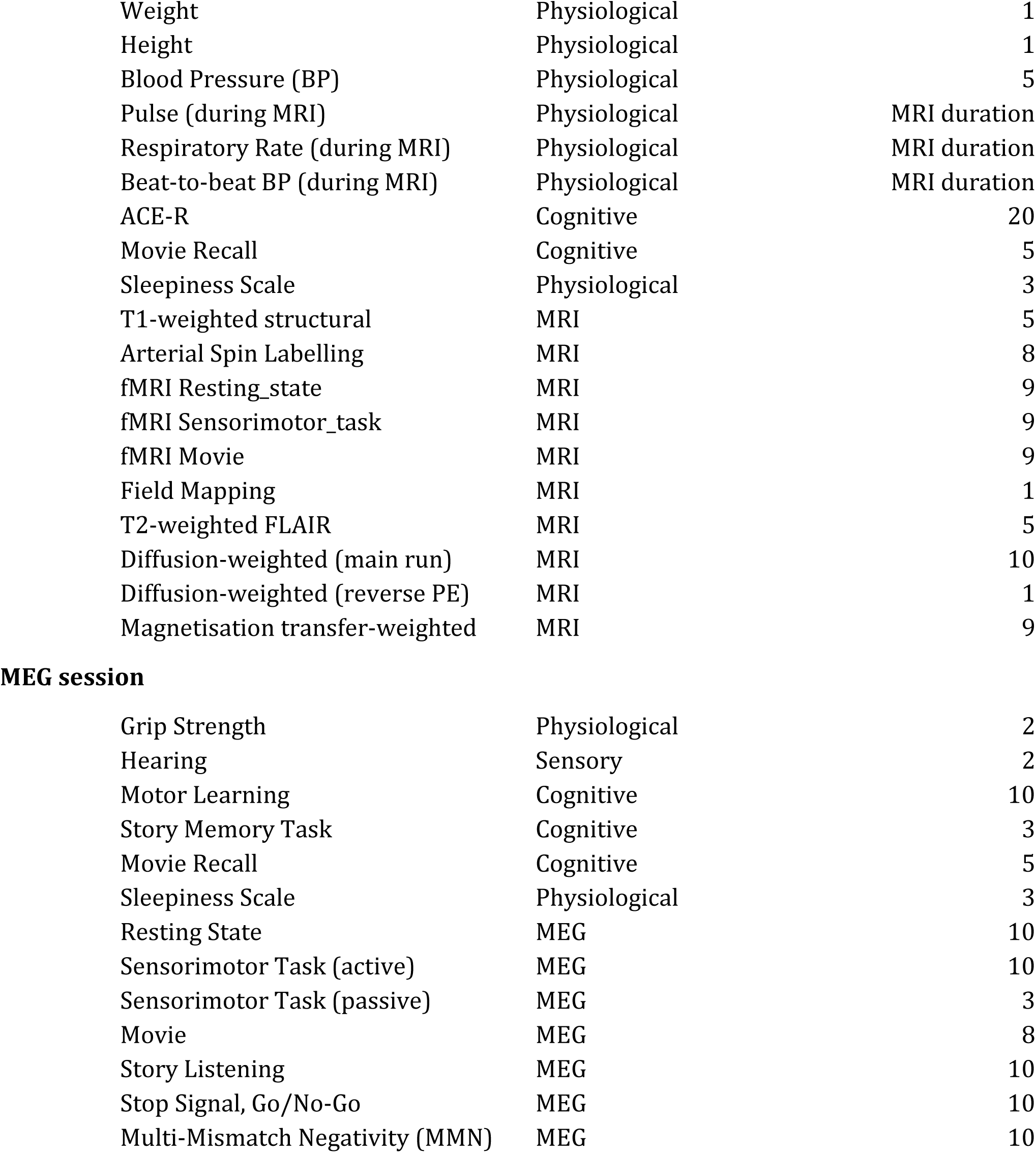
Phase 5 (Rescan) session content and timing, listed by category.

| Session | Measure | Modality | ~ Time (mins) |
| --- | --- | --- | --- |
| **Online** |  |  |  |
|  | **Core** |  |  |
|  | The Science of Ageing Questionnaire | Questionnaire | 20 |
|  | The Cattell Task | Cognitive | 20 |
|  | Spot the Word Task | Cognitive | 5 |
|  | Reaction Time Task | Cognitive | 8 |
|  | Digit-Letter Substitution Task | Cognitive | 2 |
|  | **Extended** |  |  |
|  | Visual Short-Term Memory Task | Cognitive | 20 |
|  | Spatial Memory Task (4MT) | Cognitive | 10 |
|  | Visual Long-Term Memory Task | Cognitive | 15 |
|  | Listen-Up Task | Cognitive | 10 |
|  | Word-Meaning Definitions Task | Cognitive | 12 |
|  | Predictive Inference Task | Cognitive | 15 |
|  | Sea Hero Quest | Cognitive | 10 |
| **MRI session** |  |  |  |
|  | Vision | Sensory | 2 |
|  | Hearing | Sensory | 2 |

|  |  |  |
| --- | --- | --- |
| Weight | Physiological | 1 |
| Height | Physiological | 1 |
| Blood Pressure (BP) | Physiological | 5 |
| Pulse (during MRI) | Physiological | MRI duration |
| Respiratory Rate (during MRI) | Physiological | MRI duration |
| Beat-to-beat BP (during MRI) | Physiological | MRI duration |
| ACE-R | Cognitive | 20 |
| Movie Recall | Cognitive | 5 |
| Sleepiness Scale | Physiological | 3 |
| T1-weighted structural | MRI | 5 |
| Arterial Spin Labelling | MRI | 8 |
| fMRI Resting_state | MRI | 9 |
| fMRI Sensorimotor_task | MRI | 9 |
| fMRI Movie | MRI | 9 |
| Field Mapping | MRI | 1 |
| T2-weighted FLAIR | MRI | 5 |
| Diffusion-weighted (main run) | MRI | 10 |
| Diffusion-weighted (reverse PE) | MRI | 1 |
| Magnetisation transfer-weighted | MRI | 9 |

MEG session
|  |  |  |
| --- | --- | --- |
| Grip Strength | Physiological | 2 |
| Hearing | Sensory | 2 |
| Motor Learning | Cognitive | 10 |
| Story Memory Task | Cognitive | 3 |
| Movie Recall | Cognitive | 5 |
| Sleepiness Scale | Physiological | 3 |
| Resting State | MEG | 10 |
| Sensorimotor Task (active) | MEG | 10 |
| Sensorimotor Task (passive) | MEG | 3 |
| Movie | MEG | 8 |
| Story Listening | MEG | 10 |
| Stop Signal, Go/No-Go | MEG | 10 |
| Multi-Mismatch Negativity (MMN) | MEG | 10 |

Participants were reimbursed £30 per session, with payment made after completing both sessions, plus an additional £3 per session for travel expenses.

### Core Tasks

The Science of Ageing Questionnaire covered changes since last seen in core demographics, health, memory (10MQ), current psychological health (assessed using the Hospital Anxiety and Depression Scale, HADS), sleep, women’s health, physical activity and COVID.

The Cattell Test, Spot-the-Word Task, Reaction Time Task, and Digit-Letter Substitution Task were identical to those used in Phase 4 and are described in detail in that section.

All participants with access to a computer completed the questionnaire and the four core online tests. For 21 participants without access to a computer, a paper version of the questionnaire was provided. Except for one housebound participant who did not complete the online tests, the remaining 20 participants completed the same online tests in-lab at the CBU, with a researcher present to assist with any technical difficulties.

### Extended Tasks

Phase 5 included the following online optional tasks:

1. Visual Short-Term Memory Task, which uses a continuous report paradigm to assess capacity and precision of visual short-term memory representations, as well as metacognitive awareness of recall accuracy. Based on a shortened version of the task performed in Phase 2 and reported in Mitchell, CamCAN & Cusack (2018).
2. The 4 Mountains Test (4MT) of spatial memory, which evaluates the ability to recall the topographical arrangement of four mountains within a computer-generated landscape using a delayed match-to-sample paradigm (Chan et al., 2016).
3. The Visual Long-Term Memory Task, a variant of the Mnemonic Similarity Task, which is believed to be a hippocampal-sensitive object recognition test that emphasizes pattern separation. First, participants classify how pleasant they find a series of everyday objects and animals on a scale of 1 to 5. Then, after a numerical distractor task, there is a surprise recognition test where participants must label each picture as ‘old,’ ‘similar,’ or ‘new’ (Stark et al., 2019).
4. The “Listen-Up” Task, which assesses phonological short-term memory using an automated audio-morphing method to create ambiguous syllables between pairs of words (e.g., “sheet”– “sheep”). The task is to decide which of two spoken options best resembles the test word that is initially presented (Smith et al., 2024).
5. A Word-Meaning Definitions Task, which uses a four alternative force choice task to test spoken language comprehension (modified from MacGregor et al., 2020). Participants listen to sentences with an ambiguous word that subsequently disambiguates to an unexpected meaning (e.g., “The woman disliked the mouse because of the noise it made when it clicked”). After hearing each sentence, listeners hear the ambiguous word in isolation and define the word meaning as used in the sentence by selecting the best synonym or definition from four options.
6. Predictive Inference Task, which assesses learning and belief updating. Participants estimate the obscured location of a helicopter dropping bags of coins, which they catch by positioning a bucket based on prior drops, despite visual obstructions by clouds (Nassar et al., 2016).
7. Sea Hero Quest. This optional task involved downloading the SeaHeroQuest app (https://www.alzheimersresearchuk.org/research/for-researchers/resources-and-information/sea-hero-quest/) onto their mobile phones and entering an ID that could be mapped to their CamCAN identifier. This app tests people’s ability to navigate novel mazes, and has previously been run on millions of people to investigate effects of age, sex, geography on spatial memory, and as a possible early marker of dementia (Spiers, Coutrot & Hornberger, 2023).

### In-lab physiological and cognitive measures

#### ACE-R

Before an MRI session, participants were administered the Addenbrooke’s Cognitive Examination – Revised (ACE-R) (Mioshi et al., 2006), which includes the Mini-Mental State Examination (MMSE) (Folstein et al., 1975). Different versions were used across Phase 1 (Final Revised Version A) and Phase 5 (Final Revised Version B) that had different memory components to avoid practice effects.

#### Physiological measures

To assess core physiological aspects of cardiovascular health, height, weight and blood pressure were measured (as in Phase 2). Height was recorded using a stadiometer (Seca 2017), and weight was measured with calibrated electronic scales (Seca 875). Blood pressure was measured with an oscillometric brachial cuff digital blood pressure monitor (AD Medical UM-211) after being seated for approximately 30 minutes. Three blood pressure measurements were taken, 2-3 minutes apart, to ensure reliability.

During the MRI session, pulse and respiration rate were measured using a standard Siemens photoplethysmography (pulse-oximeter) and respiratory belt. The pulse-oximeter was placed on the left index finger, and the sampling frequency for both pulse-oximeter and respiratory data was 50 Hz. Beat-to-beat non-invasive blood pressure was recorded using the CareTaker 4 device (BIOPAC part #NIBP-A-MRI), which used a finger cuff attached to the middle phalanx of the left hand. This estimated beat-to-beat blood pressure from pulse decomposition analysis, validated against arterial line measurements (Gratz et al., 2021; Kwon et al., 2022). Additionally, cardiac activity data were acquired using bipolar ECG while acquiring the MEG data, where ECG data were sampled at 1 kHz.

#### Grip Strength

In line with other ageing studies (e.g. Bennett et al., 2005), we assessed each participant’s grip strength. We used the Baseline BIMS Digital Clinic Model Grip Dynamometer. Participants held the device vertically with the upper arm against the torso, elbow 90 degrees, legs uncrossed.

They were asked to squeeze as hard as they could for 5 seconds. This was repeated three times per hand, with a 15-second rest between repetitions. The outcome measures for each repetition were mean grip force (in Newtons), grip force standard deviation, and grip force coefficient of variance.

#### Motor Learning Task

This task was designed to closely match the paradigm used in Phase 2 and described in Wolpe et al. (2020). Participants used a stylus pen to control a red cursor on a digitizing touch pad, with its position displayed on a computer monitor via a semi-reflective mirror, creating the illusion that the cursor was on the touch pad’s surface. The task required participants to move the cursor from a central home position to one of four possible target locations (0°, 90°, 180°, 270°) presented in a pseudo-random order. Successful target hits triggered a burst animation and a tone, while errors (failure to initiate movement within 1 second or reach the target within 800 ms) resulted in an error tone and a “Too slow” message. In contrast to the original task, the following conditions were included: 1) Baseline (practice) reaches with cursor visual feedback (2 cycles of 4 trials each); 2) Baseline reaches with cursor visual feedback (2 cycles); 3) Baseline reaches without cursor visual feedback (6 cycles); 4) Baseline reaches with cursor visual feedback (2 cycles); 5) Reaches with a 30-degree visuomotor rotation with cursor feedback (30 cycles); 6) After-effect measure: baseline reaches without cursor visual feedback (6 cycles). The main outcome measures included total adaptation and adaptation rate from condition (5), as described in Wolpe et al (2020).

#### Story memory task

This task was closely similar to the one used in Phase 4, with two key differences. First, story B from the Wechsler Memory Scale III (Wechsler et al., 1997) was used. Second, instead of reading the passage themselves, participants listened as the researcher read it aloud and were then asked to verbally recall the story (as was done in Phase 1). During recall, the researcher marked correct responses, and the response was audio-recorded for scoring in case of ambiguities. The 15–30 minute interval before the delayed recall test was filled with the Motor Learning Task and Grip Strength Task.

#### Sensory Assessment

Before an MRI session, participants were screened for vision and hearing. Vision was measured with the Snellen test, performed with corrected vision (i.e., glasses or contact lenses) if necessary. Appropriate vision correction was provided for the MRI and MEG scanners if needed.

Hearing was measured with the Siemens HearCheck Screener. Participants removed hearing aids before the hearing check and during the scan. Participants heard three sound intensity levels (75 dB, 55 dB, and 35 dB SPL) at two frequencies (1000 Hz and 3000 Hz). The outcome measure was the number of heard tones at each frequency in each ear. The test was performed without hearing correction to reflect participants’ natural hearing state in the scanner. Before the MEG session, participants completed an additional hearing check, where they were instructed to press a button only when they heard a sound. The outcome measure was the highest detectable dB level for each ear. In the MRI session, the sound level was adjusted based on individual hearing ability. However, in the MEG session, the sound level could not be adjusted. If participants experienced difficulty in hearing the stimulus, the researcher noted this for analysis.

### MRI session

MRI data were collected during a 90-minute session using a 3T Siemens Prisma scanner with a 32-channel head coil. Key sequence parameters are summarised below, with full details available at: https://cam-can.mrc-cbu.cam.ac.uk/materials/. The different MRI sequences are reported below in their order of acquisition.

Note that the MRI scanner used for Phase 2 and 3 was a Siemens Tim Trio, upgraded to the Prisma in December 2014.

#### T1-weighted structural image

A 3D T1-weighted structural image was acquired using a Magnetization Prepared RApid Gradient Echo (MPRAGE) sequence with the following key parameters: Repetition Time (TR) = 2250 ms, Echo Time (TE) = 3.02 ms, Inversion time (TI) = 900ms, Flip Angle (FA) = 9 degrees, Field of View (FOV) = 256 mm × 256 mm × 192 mm, voxel size = 1mm isotropic, GRAPPA acceleration factor = 2, Acquisition Time (TA) = 4 min 32 sec. This sequence is commonly used for estimating grey matter volume.

#### T2-weighted FLAIR structural image

A 2D T2-weighted Fluid Attenuated Inversion Recovery (FLAIR) image was acquired with the following parameters: TR = 9000 ms, TE =100 ms, TI =2500 ms, FA =150 degrees, FOV = 220 mm × 200 mm × 130 mm, voxel-size = 0.9 mm × 0.9 mm × 4 mm, 30% interslice gap, TA = 4 min 32 sec. This sequence is commonly used to assess white matter integrity.

#### Arterial Spin Labelling (pASL)

A pulsed Arterial Spin Labelling (pASL) sequence was acquired using perfusion mode flow- sensitive alternating inversion recovery (FAIR QII, Wong et al., 1998) with the following key parameters: TR = 4000 ms, TE = 13.22 ms, TI = 2000ms, bolus (tag) duration = 800ms, FA = 130 degrees, FOV = 240 mm × 240 mm × 144 mm, voxel size = 1.9 mm x 1.9 mm x 4.5 mm, number of slices = 32, 3D gradient-and spin-echo (GRASE) readout, Turbo factor = 32, EPI factor = 15, TA = 5 min 24 sec, including 10 label-control pairs and an M0 image. Additional three post-labelling delays were acquired: 1000ms, 2000ms, and 5000ms. This sequence is commonly used to estimate cerebral blood flow, which is closely linked to neurovascular health.

#### Resting State fMRI

T2*-weighted functional MRI (fMRI) data were acquired while participants rested with their eyes closed, as in Phase 2. Blood Oxygenation-Level Dependent (BOLD) contrast was measured using a Gradient-Echo Echo-Planar Imaging (GE-EPI) sequence. A total of 261 volumes were acquired, each containing 32 axial slices (acquired in descending order), with a slice thickness of 3 mm and a 25% interslice gap, to enable whole brain coverage including cerebellum. Key acquisition parameters: TR = 1990 ms; TE = 30 ms; FA = 78 degrees; FOV =192 mm × 192 mm; voxel-size = 3 x 3 x 3.75 mm, TA = 8 min 45 sec. This sequence is commonly used to estimate functional connectivity.

#### Sensorimotor task

To assess sensorimotor activation, fMRI data were acquired while participants performed a simple audio-visual sensorimotor task with randomly-jittered trials. Scanning parameters were the same as for the Resting State scan. This task was also used in Phase 2 and MEG (see below) and is summarised here.

Participants responded to 120 “bimodal” trials, in which two visual checkerboards (left and right of fixation) and a binaural auditory tone were presented simultaneously, cuing a motor response with the right index finger on a keypad. To ensure attention to both visual and auditory modalities, an additional 8 “unimodal” trials (four visual-only and four auditory-only) were intermixed, to which they were instructed not to respond.

The visual stimuli were displayed for 34ms, while the auditory tones were presented for 300ms. The tones were randomly selected from three frequencies: 300, 600 and 1200Hz. The minimal stimulus onset asynchrony (SOA) was 2s, but the actual SOAs were jittered by random intermixing null events, based on a second-order m-sequence optimised for estimating the haemodynamic response function (HRF). Additionally, a random jitter of 100-300ms was included to avoid aliasing of ongoing alpha rhythms (primarily for the MEG version of the task). Participants had 1500ms to respond. Before the main experiment, participants completed 5 practice trials, consisting of 3 bimodal trials (one for each 300, 600 and 1200Hz tone), one auditory-only and one visual-only trial. The behavioural outcome measures were the number of correct responses and the mean and standard deviation of the RTs.

#### Movie watching

To assess stimulus-driven (active) aspects of functional connectivity, participants were scanned while watching an excerpt of a compelling but unfamiliar film. Scanning parameters were the same as for the Resting State scan and a total of 249 volumes were collected. A similar movie was used in Phase 2 (and in MEG below). Participants watched an engaging clip from the black-and- white television drama “The Night the World Ended” by Alfred Hitchcock. The original 30-minute film was edited to a running time of 8 minutes and 8 seconds, while maintaining the plot. We refer to this clip as ”Last Day”. Participants were told that they should pay attention because they would be asked some questions about the movie later (see movie recall below). The movies themselves are available on request, subject to a research-only usage agreement.

#### Field maps

To measure magnetic field inhomogeneities, in order to unwarp the fMRI data above, an SPGR gradient-echo sequence was used with the following parameters: TR = 400 ms, with two TEs (5.19 ms and 7.65 ms), FA = 60 degrees, 32 slices with 3 mm thickness with a 25% interslice gap, FOV = 192 mm x 192 mm, voxel sixe = 3 mm isotropic, TA = 54 sec.

#### Diffusion-Weighted Images

Diffusion-Weighted Images (DWIs) were acquired with a twice-refocused spin-echo sequence, with 30 diffusion gradient directions for each of two b-values: 1000 and 2000 s/mm^2^, plus three images acquired with a b-value of 0. These parameters are optimised for estimation of the diffusion kurtosis tensor and associated scalar metrics, as well as the traditional diffusion tensor. Other parameters were: TR = 9100 ms, TE = 104 ms, voxel size =2 mm isotropic, FOV =192 mm × 192 mm, 66 axial slices, number of averages = 1, phase-encode direction anterior to posterior (AP); TA = 10 min 2 sec. A single volume with a b=0 s/mm^2^ was also acquired with reversed phase-encode direction (PA). DWI can be used to measure white-matter tracts.

#### Magnetisation-weighted Images

A baseline proton density (PD)-weighted image was acquired using a Spoiled Gradient (SPGR) sequence with the following parameters: TR = 50 ms, TE =5 ms, FA = 12 degree, FOV =210 mm × 210 mm; voxel-size =1.6 mm × 1.6 mm; bandwidth =190Hz/px, TA = 4 min and 19. A magnetisation transfer-weighted image was obtained with the same sequence as above, but including a magnetisation prepared pulse. The ratio of these images (magnetisation transfer ratio, MTR) is proposed to be sensitive to myelin density.

### Post-MRI assessments

#### Sleepiness Scale

The Karolinska Sleepiness Scale (Version B) (Miley et al., 2016) was administered after each of the three fMRI runs above. The scale consists of nine points, each corresponding to a verbal description of arousal state.

#### Movie recall

Recall of details of the movie was completed on a computer in the presence of a researcher. The test started with two pictures from the movie to cue their memory, plus two short sentences about the main characters and location. The name of the main characters were provided at the start to ensure the following questions were interpreted appropriately. After the memory cue, participants were asked if they had seen the movie before the scanning session. Then 42 open- ended questions were displayed on the screen one by one in a self-paced manner, to which the participants gave a verbal answer. The questions asked about specific details from the movie in the same chronological order as the plot of the movie was encoded (e.g., “What drink does the older gentlemen (Johnny) want from Mr Halloran?”). The researcher scored the answer immediately with three ratings: correct; incorrect; or do not remember. Upon entering this rating, the next question appeared. All answers were additionally recorded with a voice recorder. Participants were told not to worry if they could not remember everything, and encouraged to guess even if they were unsure. A list of all questions and annotations for event boundaries (from an independent sample of viewers) are available here: https://cam-can.mrc-cbu.cam.ac.uk/materials/.

### MEG session

MEG data were acquired while participants were seated inside a 306-channel Triux Neo system (MEGIN) consisting of 204 planar gradiometers and 102 magnetometers. Data were sampled at 1kHz with a band-pass filter of 0.03-330 Hz. Head position within the MEG helmet was estimated continuously using five Head-Position Indicator (HPI) coils to allow for offline correction of head motion. Two pairs of bipolar electrodes were applied beforehand to record vertical and horizontal electrooculogram (EOG) signals, along with one pair of bipolar electrodes across the chest to record the electrocardiogram (ECG) signal. Auditory stimuli were presented binaurally via plastic tubes and earpieces, while visual stimuli were projected onto a screen in front of participants.

Note that the scanner used to collect MEG data for Phases 2 and 3 was an Elekta Vectorview system, which was upgraded in March 2020 to the MEGIN Triux Neo system used in Phase 5. This involved improved electronics, but the basic sensor configuration of magnetometers and gradiometers was unchanged.

The different MEG scans are reported below in their order of acquisition.

#### Resting State

As with the fMRI resting-state scan, eyes-closed resting-state MEG data were collected for approximately 9 minutes.

#### Sensorimotor task

MEG data were recorded while participants performed the Sensorimotor Task, as described above. Following this, an additional “passive” version was conducted, in which 120 unimodal stimuli (either visual or auditory) were presented every 1000 ms, requiring no motor response. As in Phase 2, this condition was included to aid in the separation of visual and auditory sources in the MEG data.

#### Movie

As with fMRI movie-watching, MEG data were recorded while participants watched an engaging clip from the black-and-white television drama “One Grave Too Many” by Alfred Hitchcock. The original 30-minute film was edited to 8 minutes and 9 seconds, while maintaining the plot. We refer to this clip as ”I’m Not Dead”.

#### Story Listening

To assess natural language comprehension, participants listened to 10 minutes of a story from Lewis Carroll’s ‘Alice in Wonderland’, divided in two 5-minute blocks. After each block, participants answered 5 multiple-choice questions on the content of the story, to encourage and assess alertness.

#### Stop-Signal, Go/No-Go

A task similar to the online version used in Phase 4 was administered, except restricted to 83 “Go” left trials, 77 “Go” right trials, 5 “No-Go” left trials, 15 “No-Go” right trials, 17 “Stop-Signal” left trials and 23 “Stop-Signal” right trials. After a one-minute practice, the main task lasted 7 minutes.

#### Multi-Mismatch Negativity

This task measured neural responses to unexpected auditory events, which have been shown to depend on fronto-temporal interactions. The task was adapted from the multi-feature “Optimum- 1” paradigm (Näätänen et al., 2004) as transcribed to our MEG by Hughes and Rowe (2013). In two, 5-minute blocks, harmonic tones were presented every 500 milliseconds while participants watched a silent natural history film. Standard tones had a 75 ms duration with three sinusoidal partials of 500, 1000 and 1500 Hz. Five types of deviant tones differed from the standard in either frequency (550, 1100, and 1650 Hz), intensity (+/- 6dB), duration (25 vs. 75 ms), sound location (left or right rather than bilateral), or by a silent gap of 25 ms in the middle of the tone.

Each block started with 15 standard tones, after which deviant tones were interspersed among the standard tones. The order of deviant tones was permuted, such that in a sequence of 10 tones, each deviant was presented once, and the same deviant type was never presented consecutively. In total, 600 standard tones and 600 deviant tones were presented. This task was very similar to that used in Phase 3 (600 versus 900 tones)

### Post-MEG assessments

#### Sleepiness Scale

The Karolinska Sleepiness Scale (see above) was administered after the resting state, Sensorimotor Task, movie, and Multi-Mismatch Negativity runs.

#### Movie recall

The MEG movie recall task followed the same structure as the MRI movie recall but included 38 questions instead of 42.

## Derived Variables

Some important summary variables were derived from questionnaire responses, as detailed below, and are available on https://cam-can.mrc-cbu.cam.ac.uk/materials/.

### Years of Education

Foremost was the total number of years in education. A base number of years was assumed according to year of birth, owing to changes in UK educational policy. Those born before 1 September 1933 were assumed to have a base of 9 years, whereas those born between this date and 1 September 1956 were assumed to have a base of 10 years, owing to the change in mandatory schooling from 1 September 1947 onwards (the “Education Act of 1944”). Those born after 1 September 1956 were assumed to have a base of 11 years, owing to a further increase in mandatory schooling following the “Raising of the School leaving Age” (ROSLA) Act in 1972. Then years on top of this base score were determined by qualifications. Our original Phase 1 questionnaire asked for the highest standard UK educational qualification (e.g., none, GCSE/CSE/O-level/NVQ/HND/HNC, A-level, university degree), as well as professional qualifications (e.g., teaching, nursing). For those with GCSE-equivalents, 2 years were added for those born before 1933, 1 year was added for those born between 1933 and 1956 (since after ROSLA, the extra year counted as the first year of the CSE/O-levels), and no years were added for those born after 1956 (as GCSE-equivalents were part of the last two years of mandatory education). A further 2 years was added for A-levels, and a further 3 years for university degrees. Then a further 2 years were added for each professional qualification. Thus, for example, a person born after 1956 would be assumed to have 11 years of education if they responded “none” or their highest qualification was GCSE or equivalent; 13 years if their highest qualification was A-levels; 16 years if their highest qualification was a degree; or 18 years if they had a degree and another professional qualification.

### Physical Activity

Phase 1 administered the EPIC-Norfolk Physical Activity Questionnaire (EPAQ; Wareham et al., 2002). This was too long to repeat fully in Phases 4 and 5, so a reduced set of questions were asked about vigorous, moderate, light and sedentary activities. For each of these four types, participants were asked: 1) how many days within the last 7 included such activity, and 2) how many hours and minutes were spent on a typical one of those days. Note that, owing to an error, the number of minutes of “light” activity was not recorded in Phase 5, so was set to the mean of all light activity minutes from Phase 4. Also, “sedentary” activity in Phases 4 and 5 was restricted to a question about time “sitting down”, which may only be a subset of what people interpreted as sedentary in Phase 1.

By combining across these four categories, one can estimate the weekly Physical Activity Energy Expenditure (PAEE) per person. However, because the precise formula for PAEE cannot be applied to this subset of questions, the weights for each of these four categories were estimated by a linear, least-squares fit of Phase 1 PAEE values by this subset of questions in Phase 1. These weights were then applied to the Phase 4 and 5 values to estimate PAEE.

### Summary of Longitudinal Data

Table 3 shows a summary of data with more than one assessment across all 5 Phases.

**Table 3.** Data availability summary across Phases.

| Measure | Phase 1 | Phase 2 | Phase 3 | Phase 4 | Phase 5 |
| --- | --- | --- | --- | --- | --- |
| **Questionnaire** | Y | - | - | Y | Y |
| **Cognitive measures** |  |  |  |  |  |
| ACE-R | Y | Y (Arm2) | MMSE only | - | Y |
| Cattell | - | Y | MRI version | Y | Y |
| Spot the Word Task | Y | Y (Arm2) | - | Y | Y |
| Reaction Time | Y | - | MRI version | Y (opt) | Y |
| Verbal Memory | Y | - | - | Y (opt) | Y |
| Digit-Symbol Substitution | - | Y (Arm2) | - | Y (opt) | Y |
| Visual Short Term Memory | - | Y | MRI version | - | Y (opt) |
| Motor learning | - | Y | - | - | Y (opt) |
| Stop-Signal, Go/No-Go | - | - | MEG version | Y | MEG version |
| **Physiological measures** |  |  |  |  |  |
| Blood Pressure | - | Y | Y | - | Y |
| ECG | - | Y | Y | - | Y |
| Height | - | Y | Y | - | Y |
| Weight | - | Y | Y | - | Y |
| Grip Strength | - | - | - | - | Y |
| Motor Learning | - | Y (Arm1) | - | - | Y |
| Hearing/Vision | Y | Y | Y | - | Y |
| **MRI** |  |  |  |  |  |
| T1-weighted structural | - | Y | Y | - | Y |
| Arterial Spin Labelling | - | Y (Arm2) | Y | - | Y |
| Resting State | - | Y | Y | - | Y |
| Sensorimotor Task (active) | - | Y (Arm1) | - | - | Y |
| Movie | - | Y | - | - | Y |
| Field Mapping | - | Y | Y | - | Y |
| T2-weighted SPACE/FLAIR | - | Y (Arm1) | Y | - | Y |
| Diffusion-weighted (main run) | - | Y | - | - | Y |
| Magnetisation transfer-weighted | - | Y | - | - | Y |
| **MEG** |  |  |  |  |  |
| Resting State | - | Y | Y | - | Y |
| Sensorimotor Task (active) | - | Y | - | - | Y |
| Sensorimotor Task (passive) | - | Y | - | - | Y |
| Movie | - | - | - | - | Y |
| Stop-Signal, Go/No-Go | - | - | Y | - | Y |
| Multi-Mismatch Negativity | - | Y (Arm2) | Y | - | Y |

Note that sometimes the test format (online/paper, at-home/in-lab) varied across Phases, and MRI versions differed from main versions of task. Opt = optional.

## Discussion

The continuation of the CamCAN study presents a rare opportunity to study long-term changes in neurocognitive function across the adult lifespan, with repeated, multi-modal brain assessments over 12 years being particularly rare for a population-derived healthy cohort (and currently unique for MEG). These longitudinal data will allow tracking of individual changes, controlling for the generation differences that confound cross-sectional studies of ageing. Together with other longitudinal cohorts around the world, the CamCAN data allow a rich and multi-faceted characterisation of age-related changes in brain and cognition, along with lifestyle and demographic predictors.

## Data Availability

All data produced in the present study will be available upon reasonable request to the authors, as we have done with previous waves of CamCAN.

https://camcan-archive.mrc-cbu.cam.ac.uk/dataaccess

## Competing interests

The authors declare that they have no competing interests.

## Authors’ contributions

The protocol was designed by RH, KT, KC, NW & JR, with individual tasks contributed by DJM, NW, KT and AA. Recruitment was coordinated by TE. Data were collected by LB, ID & WD. Data were collated by AA. ID prepared the manuscript. All authors contributed text to the manuscript and provided feedback. All authors approved the last version of the manuscript.

## Authors’ information

CamCAN co-author includes the following: Project principal personnel: Richard N Henson, Lorraine K Tyler, Kamen A Tsvetanov, Carol Brayne, Edward T Bullmore, Andrew C Calder, Rhodri Cusack, Tim Dalgleish, John Duncan, Fiona E Matthews, William D Marslen-Wilson, James B Rowe, Meredith A Shafto, Marta Correia; Research Associates: Karen Campbell, Teresa Cheung, Simon Davis, Linda Geerligs, Rogier Kievit, Anna McCarrey, Abdur Mustafa, Darren Price, David Samu, Jason R Taylor, Matthias Treder, Janna van Belle, Nitin Williams, Daniel J Mitchell, Simon Fisher, Else Eising, Ethan Knights, Adam Attaheri, Dace Apsvalka, Maite Crespo-Garcia; Research Assistants: Lauren Bates, Tina Emery, Sharon Erzinçlioğlu, Andrew Gadie, Sofia Gerbase, Stanimira Georgieva, Claire Hanley, Beth Parkin, David Troy, Ina Demetriou, Will Duckett; Affiliated Personnel: Tibor Auer, Lu Gao, Emma Green, Rafael Henriques; Research Interviewers: Jodie Allen, Gillian Amery, Liana Amunts, Anne Barcroft, Amanda Castle, Cheryl Dias, Jonathan Dowrick, Melissa Fair, Hayley Fisher, Anna Goulding, Adarsh Grewal, Geoff Hale, Andrew Hilton, Frances Johnson, Patricia Johnston, Thea Kavanagh-Williamson, Magdalena Kwasniewska, Alison McMinn, Kim Norman, Jessica Penrose, Fiona Roby, Diane Rowland, John Sargeant, Maggie Squire, Beth Stevens, Aldabra Stoddart, Cheryl Stone, Tracy Thompson, Ozlem Yazlik; and administrative staff: Dan Barnes, Marie Dixon, Jaya Hillman, Joanne Mitchell, Laura Villis.

## Acknowledgements

We thank the Cam-CAN respondents and their primary care teams in Cambridge for their participation in this study, and colleagues at the MRC Cognition and Brain Sciences Unit MEG and MRI facilities for their assistance. Cam-CAN was initially supported by the Biotechnology and Biological Sciences Research Council Grant BB/H008217/1, then received further contributions from the European Union’s Horizon 2020 research and innovation programme (‘LifeBrain’, Grant Agreement No. 732592) and intramural funding to the MRC Cognition & Brain sciences Unit [SUAG/046/G101400]. K.A.T was supported by Fellowship awards from the Alzheimer’s Society (Grant Nr. 6002) and the Guarantors of Brain (G101149). J.B.R. is supported by the Wellcome Trust (220258) and the NIHR Cambridge Biomedical Research Centre (NIHR203312). For the purpose of open access, the author has applied a Creative Commons Attribution (CC BY) licence to any Author Accepted Manuscript version arising from this submission. We thank Tom Hartley, Matt Davis, Hugo Spiers, Claire Lancaster for assistance with some of the tasks.

## Footnotes

1 Note that we used the term “stage” in Shafto et al. (2014) rather than “phase”, but these two terms are inter-changeable within CamCAN.

## Notes

Funding: Cam-CAN was originally supported (2010-2015) by the Biotechnology and Biological Sciences Research Council (BBSRC) Grant BB/H008217/1. It was subsequently supported by the Medical Research Council (MRC) core Unit grant (SUAG/094 G116768) and Programme grant to R.H. (SUAG/086 G116768), with a contribution (2017-2022) from the European Union Horizon 2020 Research and Innovation Program (LifeBrain) Grant Agreement 732592.

### Competing Interest Statement

The authors have declared no competing interest.

### Funding Statement

This study was originally funded (2010-2015) by the Biotechnology and Biological Sciences Research Council (BBSRC) Grant BB/H008217/1. It was subsequently supported by the Medical Research Council (MRC) core Unit grant (SUAG/094 G116768) and Programme grant to R.H. (SUAG/086 G116768), with a contribution (2017-2022) from the European Union Horizon 2020 Research and Innovation Program (LifeBrain) Grant Agreement 732592.

### Author Declarations

Cambridgeshire 2 Research Ethics Committee, of the UK, gave ethical approval for this work (reference: 10/H0308/50, Amendment 7)

